# ACE2 gene expression and inflammatory conditions in periodontal microenvironment of COVID-19 patients with and without diabetes evaluated by qPCR

**DOI:** 10.1101/2022.03.10.22271304

**Authors:** Boy M. Bachtiar, Endang W. Bachtiar, Hari Sunarto, Yuniarti Soeroso, Benso Sulijaya, Citra Fragrantia Theodorea, Irandi Putra Pratomo, Yudhistira, Ardiana Kusumaningrum, Defi Efendi, Efa Apriyanti, Nastiti Rilo Utami, Astri Deviana, Anissa Dien Andriyani

## Abstract

**Objective:** Chronic periodontitis has been proposed to be linked to coronavirus disease (COVID-19) on the basis of its inflammation mechanism. We aimed to evaluate this association by investigating the expression of Angiotensin Converting Enzyme-2 (ACE2) in periodontal compartments, which contain dysbiosis-associated pathogenic bacteria, and how it can be directly or indirectly involved in exacerbating inflammation in periodontal tissue.

**Material and Methods:** This observational clinical study included 23 adult hospitalized patients admitted to Universitas Indonesia Hospital with PCR-confirmed COVID-19, while 6 non-COVID-19 participants come to periodontal clinic were included as control. Using real time-PCR (qPCR) and gingival crevicular fluids (GCF) samples from COVID-19 patients with and without diabetes and periodontitis, we assessed the mRNA expression of angiotensin-converting enzyme 2 (ACE2), IL-6, IL-8, complement C3, and LL-37 as well as the relative proportion of *Porphyromonas gingivalis, Fusobacterium nucleatum*, and *Veillonella parvula* to represent the dysbiosis condition in periodontal microenvironment. All analyses were performed to determine their relationship.

**Results:** *ACE2* mRNA expression was detected in the GCF of periodontitis-COVID-19 patients with and without diabetes. However, only periodontitis-COVID-19 patients with diabetes showed a positive relationship between *ACE2* expression and inflammatory conditions in the periodontal microenvironment. In addition, the interplay between pro-inflammatory cytokine (IL-6) and complement C3 could be used as a predictor of the severity of periodontal inflammation in COVID-19 patients with diabetes.

**Conclusion:** The study data show that the SARS-CoV-2 entry gene is expressed in the GCF of patients with COVID-19, and its expression correlates with inflammatory markers.

## Introduction

From a dental perspective, the outbreak of the COVID-19 pandemic poses a threat to the practice of dentistry. This is because the oral cavity should be considered a preferential route for the entry or transmission of the related-virus (SARS-CoV-2) among populations that can occur directly or indirectly [1-3], particularly in nosocomial environments at risk [4]. Recent studies have shown that COVID-19 infection and severity can be linked to the presence of periodontitis [5]. This association is based on the fact that periodontitis and COVID-19 share common risk factors that enhance chronic diseases, including diabetes, hypertension, age, sex, and genetic variants [6, 7]. Another possible explanation is that the aspiration of periodontopathogens observed in periodontitis patients could have a synergistic effect with the virus or compromise the host immune response against COVID-19 [8]. Moreover, studies have shown that in addition to a wide variety of human tissues, the genetic material of SARS-CoV-2 can be identified in periodontal tissue [9, 10], gingival crevicular fluid (GCF) [11], and saliva of infected patients [12, 13].

To enter cells, this virus requires angiotensin-converting enzyme 2 (ACE2) as the host receptor [14-17]. In oral tissues, the receptor can be detected in tongue mucosa [18] and periodontal tissue [19]. This indicates that the oral tissues are susceptible to SARS-CoV-2 infection. In addition, certain molecules (furin, cathepsin, and TMPRSS2) are involved in the entry of the virus into cells and are found in the periodontal tissue [20]. However, much debate has centered on the question of whether the periodontal microenvironment conditions may also provide favorable conditions for SARS-CoV-2.

In this study, we aimed to explore the presence of determinants of SARS-CoV-2 infection in the subgingival niche and the nature of the possible association with periodontitis. Here, we compared the abundances of selected periodontal pathogens, *P. gingivalis, F. nucleatum*, and *V. parvula*, in the subgingival microbiota. Furthermore, we analyzed the association between *P. gingivalis* proportion and the transcription levels of *ACE2*, and IL-6, IL-8, C3, and LL-37 genes in infected COVID-19 patients with and without diabetes and diagnosed with chronic periodontitis. Therefore, the clinical features of periodontal disease, diabetes, and COVID-19 are beyond the scope of our observational study. Greater knowledge of innate immunity in the periodontal compartment may provide valuable insights into the link between COVID-19, periodontal inflammation, and diabetes.

## Methods

### Patients

Before starting this study, we obtained ethical approval from the Ethical Review Committee of Medical Ethical Committee of Rumah Sakit Universitas Indonesia (RSUI) and followed the principles of the Medical Research Involving Human Subject Act (ETHICAL APPROVAL Nomor: 0042/SKPE/KKO/2021/00). All sampling methods were carried out in accordance with committee guidelines and regulations, and this study was carried out in accordance with the guidelines provided by the Strengthening the Reporting of Observational Studies in Epidemiology (STROBE) statement [21].

After a briefing on the procedure, only participants who provided written consent were included in the study. We obtained the clinical conditions of all recruited patients and the subjects needed for control from the hospital medical records. We collected information regarding age, sex, chronic medical history of comorbidities, and clinical symptoms. However, as stated above, our concern was for COVID-19 patients with comorbidities of periodontitis, with and without diabetes. Thus, we did not include these variables in the data analysis.

A total of 23 patients with periodontitis and COVID-19 staying in Rumah Sakit Universitas Indonesia (RSUI), Depok, Jawa Barat, Indonesia, were included in this study between June 15, 2021, and July 25, 2021. They were requested to participate in this study after their COVID-19 status was confirmed by nasopharyngeal swab testing using RT-PCR at least seven days prior. Among the 23 subjects, ten patients reported to have diabetes type 2. Only subjects who had no respiratory symptoms for more than 2 weeks were selected.

COVID-19 patients with diabetes were defined according to the medical report obtained from the hospital, while all COVID-19 patients were diagnosed with periodontitis (moderate to severe) according to the criteria described by the American Academy of Periodontology Classification of Periodontal Disease [22]. Periodontitis assessment and clinical evaluation were performed by a registered dentist. For controls, we used GCF collected from non-COVID-19 individuals (n = 6) who were registered in the periodontal clinic of the RSUI. Participants who were unwilling to provide written informed consent were excluded from the study. Other exclusion criteria were previous periodontal treatment within the last year, age < 18 years, use of antibiotics during the last 6 months, and pregnancy.

### Sample collection

Because it is difficult to accurately determine the periodontal sites that undergo progressive tissue breakdown, we relied on detecting the signs of tissue damage by measuring the probing pocket depth (PPD) to detect loss of attachment [23, 24]. PPD was determined as previously reported [25]. We conducted a professional procedure to collect GCF. GCF samples were collected as previously reported [26]. Briefly, the GCF samples were collected before bacterial sampling and from one diseased site with a probing depth of ≥3 mm with bleeding on probing. The collection area was isolated with cotton rolls, and supragingival plaque was carefully removed with curettes. Collection was performed with a sterile endodontic paper point by gently inserting the point to the depth of the sulcus and moving it laterally along the axis of the tooth. To obtain a sufficient amount of GCF, multiple paper points were collected from several tooth sites. Any paper points visibly contaminated with blood were discarded. Immediately after sampling, the paper points were pooled in microcentrifuge tubes containing 100 μL of phosphate buffer and RNAlater (Invitrogen, Carlsbad, CA, USA), and subsequently stored at □70°C until further analysis.

### Proportion of P. gingivalis, F. nucleatum, and V. parvula in subgingival microbiota

The levels of bacterial (*P. gingivalis, F. nucleatum*, and *V. parvula*) colonization and the total amount of bacteria in the periodontal microenvironment were determined using quantitative real time PCR (qPCR) with the primers listed in Table 1. Their relative proportions were determined relative to the total bacteria in the same oral niche.

**Table 1.**
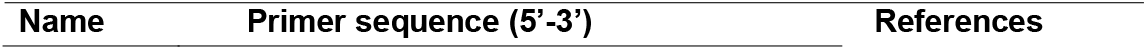
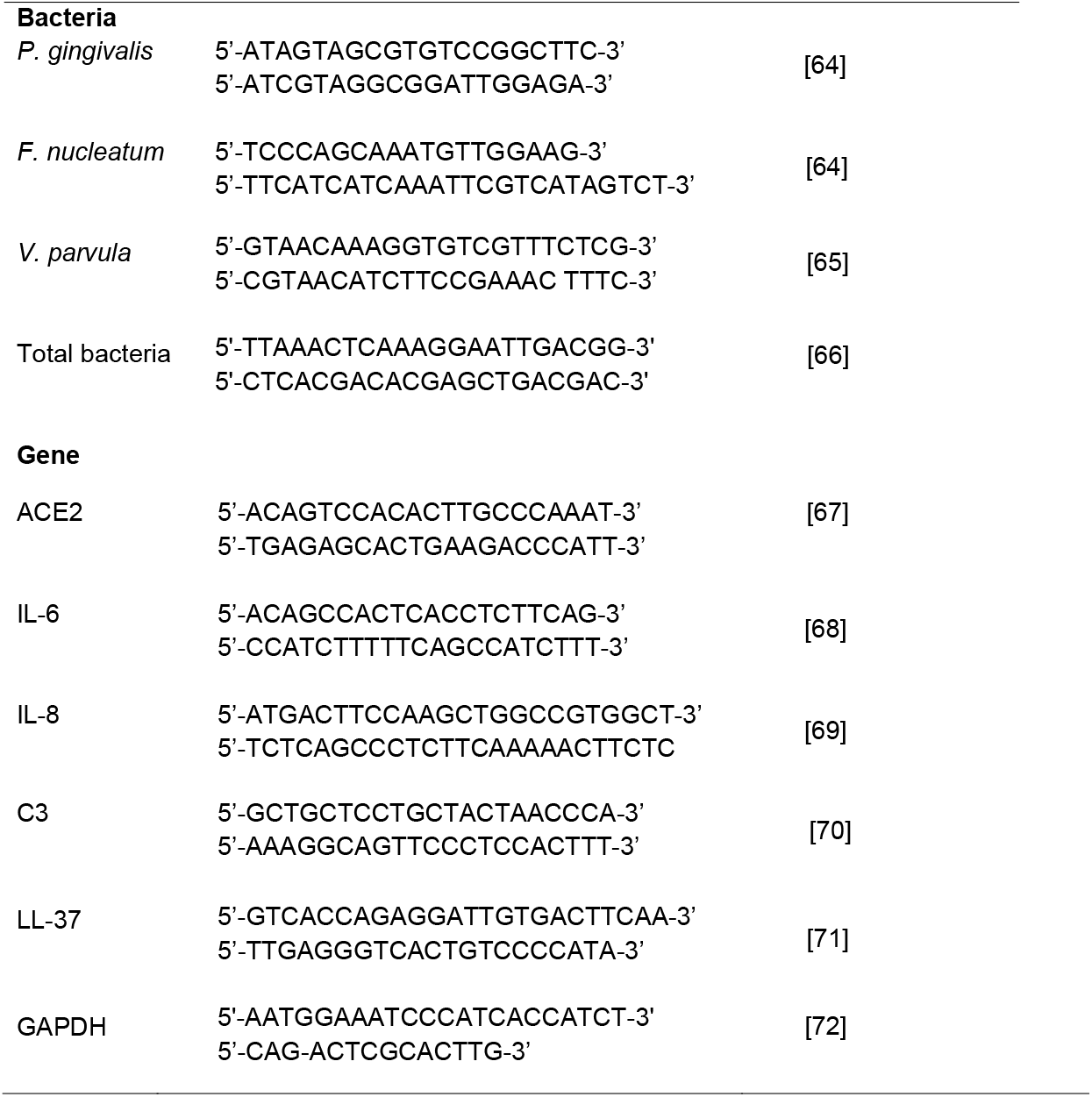
Primers used in this study

Therefore, we first extracted DNA from the GCF samples using GENEzol™ reagent (General, Ltd, New Taipei City, Taiwan), following the manufacturer’s instructions. The concentration and quality of the DNA were determined using Qubit assay reagents (Invitrogen, Carlsbad, CA, USA). After dissolving in Tris-EDTA buffer, the DNA was cooled to □20°C until further processing. Prior to PCR amplification, the amount of DNA in the samples was measured using SYBR green PCR Master Mix (Applied Biosystems) and specific primers (Table 1) using the ABI StepOnePlus Real-Time PCR system.

The qPCR reaction was carried out in a total volume of 10 μL, containing 5 μL of SYBR1 Selected Master Mix (Thermo Fisher Scientific, Waltham, USA), 2 μL of DNA template, and 1 μL of primer pair solution (300 nM/ reaction). For each run, DEPC-treated water (Thermo Fisher Scientific, Waltham, MA, USA) was used as a negative control, and melting peaks were used to determine PCR specificity. qPCR analysis was performed in triplicate using ABI StepOnePlus Real-Time PCR Master Mix (Applied Biosystems) following the manufacturer’s protocol. The thermal cycling conditions were set as follows: pre-denaturation at 95°C for 10 min, followed by 40 cycles at 95°C for 15 s, 55°C for 30 s, and an additional step at 72°C for 15 s.

The relative abundance of each bacterium was calculated using the relative calculation of the ΔΔCt method (2^-ΔΔCt^) [27]. ΔCt was calculated as the difference between the Ct value using the primers for each bacterium and the Ct value using the primers for total bacteria. ΔΔCt was defined as the difference between the ΔCt value of the patient and control subjects, where the value derived from the 2^-ΔΔCt^ shows the changes in bacterial abundance in the sample of patients relative to those of the control subjects. The 2^-ΔΔCt^ value of the control was 1.

### Quantitation of IL-6, IL-8, C3, and LL-37 transcription by qPCR

RNA was extracted from GCFs using GENEzolTM reagent (General, Ltd, New Taipei City, Taiwan), according to the manufacturer’s instructions, followed by a reverse transcription kit (Applied Biosystems). The resulting cDNA was amplified by qPCR with specific primers, as shown in Table 1, while non-transcribed RNA samples were used as controls for genomic DNA contamination. The genes studied in this investigation were angiotensin-converting enzyme 2 (ACE2), interleukin (IL-6 and IL-8, complement C3, and antimicrobial peptide LL-37). qPCR analysis was performed in triplicate on an ABI StepOnePlus Real-Time PCR System with SYBR Green PCR master mix (Applied Biosystems), according to the manufacturer’s instructions. The PCR conditions were as follows: pre-denaturation at 95°C for 5 min, followed by 40 cycles of 95°C for 10s, 60°C for 30s, and 72°C for 30s, and a final extension at 72°C for 5 min. The melting curve profile was set as follows 95°C for 15s, 60°C for 60 s, and 95°C for 15 s. In this study, gene expression was normalized to the level of d-glyceraldehyde-3 phosphate dehydrogenase (GAPDH), and the mRNA expression of each target gene in GCF from non-COVID-19 patient was set as the control. Results greater than or less than 1 were considered to indicate up- or downregulation of mRNA expression. All values obtained from the tested patient groups were standardized and compared to the control value, represented as a value of 1.

### Statistical analysis

Provided that we did not have access to clinical symptoms regarding the COVID-19 condition and blood glucose of the patients from which the oral samples were collected, the relevance of such variables in the statistical analysis could not be assessed. In this study, the relative abundance of bacteria and transcription levels of mRNA in the targeted gene were analyzed in two patient groups: periodontitis-COVID-19 with (G1) and without (G2) diabetes.

Statistical analyses were conducted using GraphPad PRISM 9.0 (GraphPad Software, San Diego, CA, USA). Data are presented as mean ± standard error (SE). Statistical significance was set at P < 0.05. For comparison between groups, P-values were determined by one-way analysis of variance (ANOVA), whereas for comparison within groups, two-tailed P-values were determined by unpaired Student’s *t*-test. For correlation analysis, Spearman’s correlation coefficient (r) with two-tailed P-values was calculated, and linear regression was used to generate the line of best fit with 95% confidence intervals. Based on the correlation results, we used receiver operating characteristic (ROC) curve analysis to determine the optimal threshold for detecting inflammation markers with high sensitivity and specificity.

## Results

This study evaluated 23 patients with confirmed COVID-19 infection admitted to Universitas Indonesia Hospital (RSUI). Six of the control subjects were those who visited the periodontal clinic in RSUI. The mean ± SD age of the 29 participants was 45.1 ± 15.37-year-old. As far as COVID-19 and diabetes cases are concerned, we found that 10 of the 23 enrolled patients were diabetic.

First, the transcription levels of *ACE2* and inflammation elements between groups were compared, followed by the calculation of the abundance of DNA load of each bacterium selected and a comparison of their proportions among groups (G1 and G2). Finally, we analyzed the correlation between independent variables observed in each group tested.

### ACE2 mRNA levels and relative abundance of P. gingivalis

To predict how susceptible cell-associated periodontal tissue is to SARS-CoV-2, we examined the mRNA expression of *ACE2* in GCF samples from G1 and G2 groups. *ACE2* mRNA expression in GCF samples from healthy (non-COVID-19) patients was included as a control. Our data showed that *ACE2* mRNA was detected in GCF samples collected from all the subjects tested (G1, G2, and control). Given that the SARS-CoV-2 receptor is highly enriched in epithelial cells of the tongue [18], we compared *ACE2* transcription levels in GCF and tongue surface (TS) swab samples. On average, the transcription level of *ACE2* was lower in the GCF than that in the tongue surface (P < 0.05). When the gene expression was compared between the G1 and G2 groups, we found that in GCF, the expression of *ACE2* was significantly higher in the G1 group (2-fold, P < 0.05) than in the G2 group, while in TS, the difference was not significant (Fig. 1A, P > 0.05). Subsequently, we found that all surveyed bacteria could be detected in the subgingival microbiota of all subjects tested. The qPCR results showed that in both groups, the proportions of *P. gingivalis* and *F. nucleatum* were comparable, whereas the proportion of *V. parvula* was significantly higher in G1 than in G2. Interestingly, in both groups, *the number of P. gingivalis* was found to be the lowest compared to the abundance of the other two species. The proportion of *F. nucleatum* and *V. parvula* was >30% and >50% higher than that of *P. gingivalis, respectively (*Fig. 1B, P < 0.05).

**Fig. 1.**
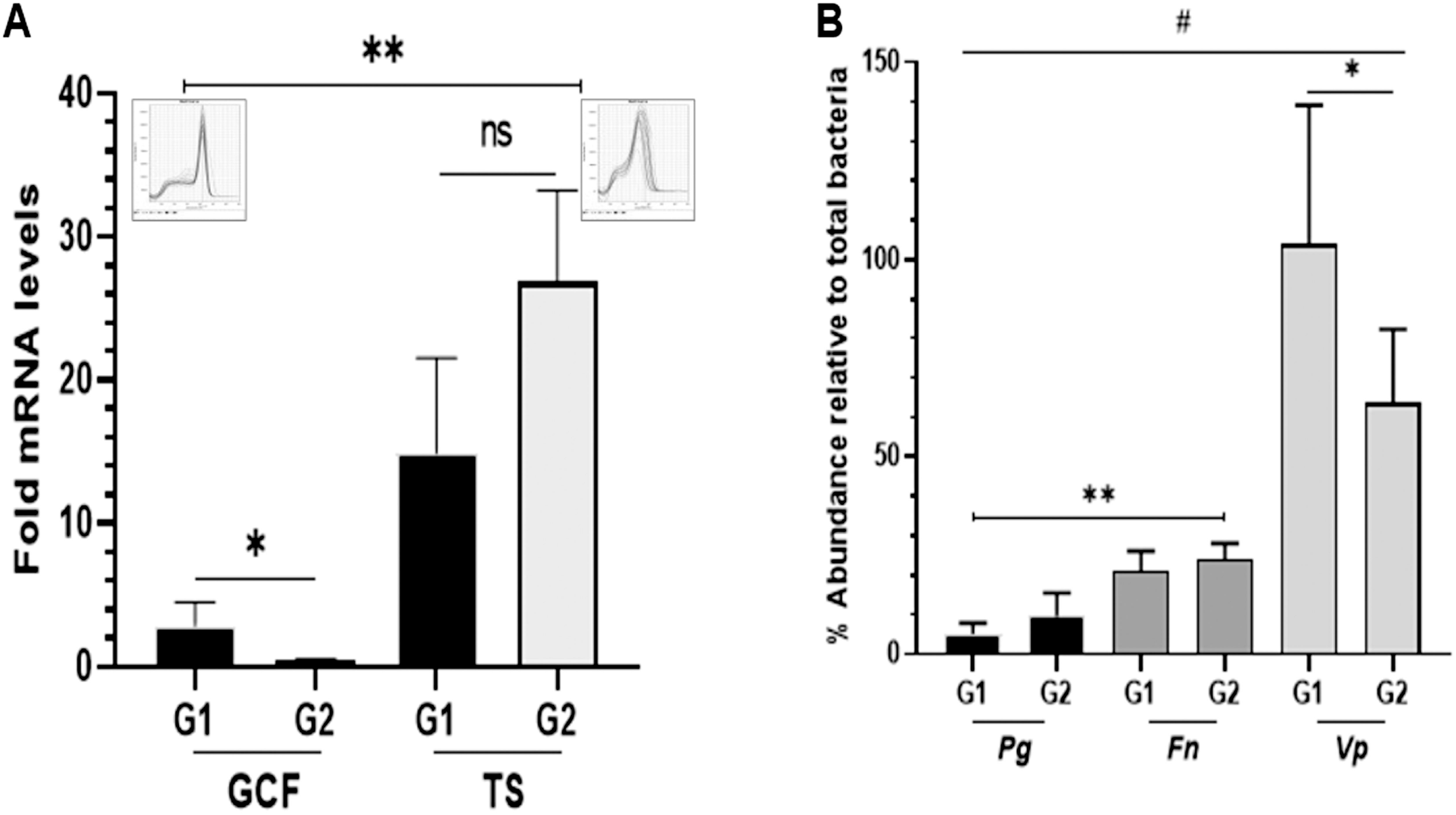
Evaluation of *ACE2* expression (A), and the proportion of *P. gingivalis* (Pg), *Fusobacterium nucleatum* (*Fn*), and *Veillonella parvula* (*Vp*) (B), in different patient groups of COVID-19. The transcription level of *ACE2* mRNA was higher in COVID-19 patients with (G1) diabetes than those without (G2) diabetes, but the reverse was found in tongue surface (TS). The proportion of *Pg* was found to be the lowest compared to the other two species (*Fn* and *Vp*). However, the relative abundance of this key stone species [43] was not significant in either patient (G1 or G2). All data are expressed as mean ± SE. * significant difference of the gene expression within group (G1 and G2). ** significant different of the gene expression between GCF and TS or between *Pg* and *Fn*. NS = not significant difference. # significant different of the bacterial abundance *Vp* and *Pg* and *Fn*. The inserts denote the melting curve of qPCR in GCF (left) and TS (right) samples.

Furthermore, a strong positive correlation was observed between the proportion of *P. gingivalis* and *F. nucleatum* in the G1 and G2 groups (r = 0.95, P = 0.0001, and r = 0.82, P = 0.0008), while moderate (r = 0.52, P = 0.01) and strong (r = 0.98, P = 0.001) correlations were observed between *P. gingivalis* and *V. parvula* in the G1 and G2 groups, respectively (Fig. 2A-D).

**Fig. 2.**
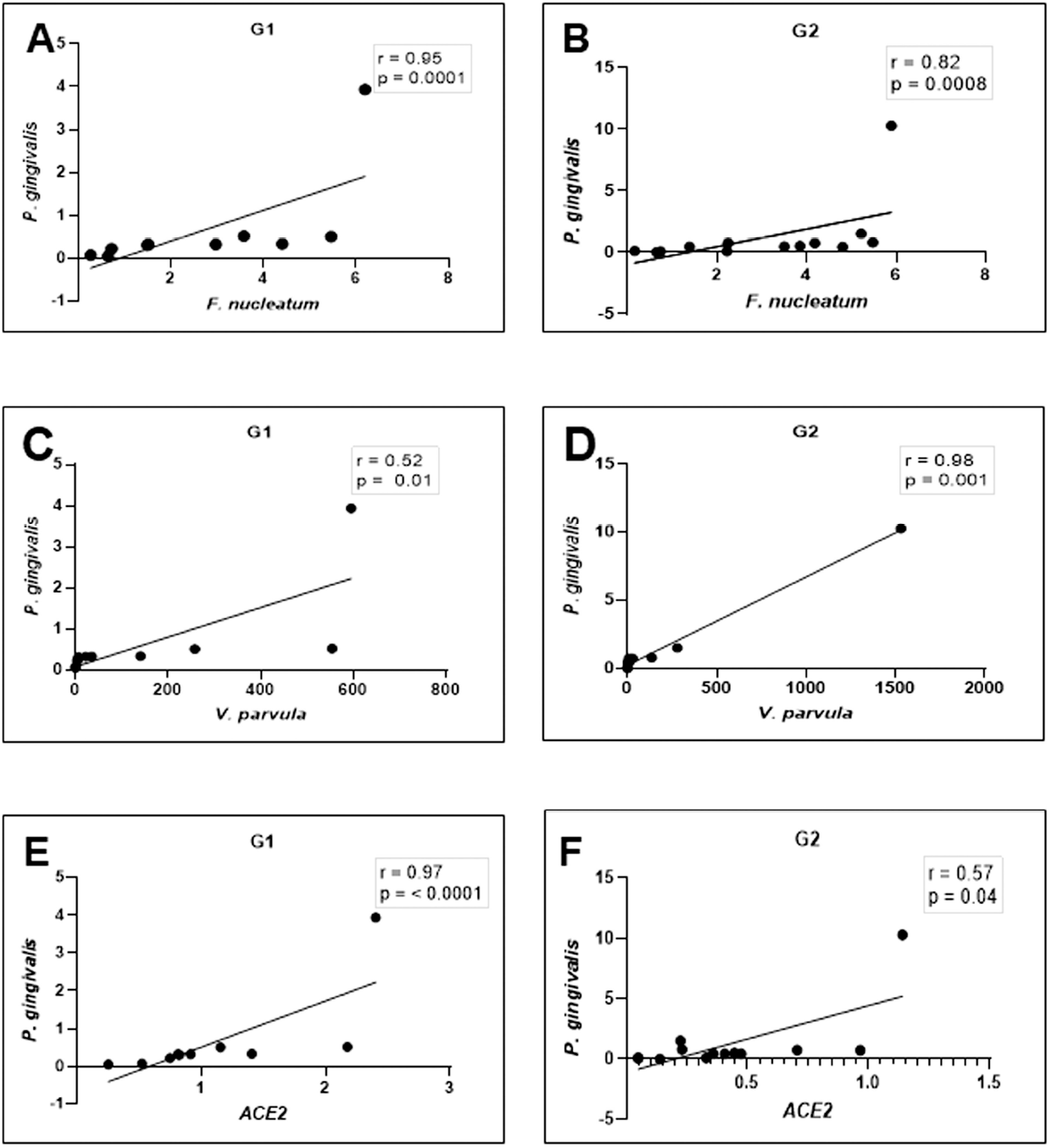
Relationship between the relative proportion of *P. gingivalis* and the two Gram-negative oral bacteria (*F. nucleatum* and *V. parvula*) (A-D), and between *P. gingivalis* abundance and *ACE2* mRNA expression levels (E and F). In either patient group (G1 or G2), the correlations were significant moderate (C and F) and strong positive (A, B, D, and E).

To further examine the relationship between *ACE2* expression and the relative abundance of the keystone pathogen (*P. gingivalis*) [28], we analyzed the correlation between the transcription level of *ACE2* and the relative proportion of the bacterium in each group studied. As shown in Fig. 2E and F, a strong and moderate linear positive correlation between the transcription of *ACE2* mRNA and the relative abundance of *P. gingivalis* was observed in groups G1 (r = 0.97, P = 0.0001) and G2 (r = 0.57, P = 0.04), respectively.

### Transcription levels of immuno-inflammatory markers (IL-6, IL-8, C3, and LL-37) and their relationship with ACE2 mRNA and relative abundance of P. gingivalis

Based on the above results, two proinflammatory cytokines (IL-6 and IL-8) and two innate response elements (C3 and LL-37) were further analyzed to investigate their relationship with *ACE2* mRNA expression and the proportion of *P. gingivalis*. We found that IL-6 was detectable in all GCF samples, whereas IL-8 was only measured in 6/10 (60%) and 10/13 (77%) of the G1 and G1 patient groups, respectively. Statistical analyses revealed that the gene expression of IL-6 was upregulated by 2-fold in the G1 group compared with mRNA expression, which showed a 1.5-fold increase in the G2 group (P < 0.001). The expression of IL-8 mRNA was found to be comparable (2-fold, P > 0.05) in both groups. In terms of the C3 and LL-37 mRNA expression levels, the detection rate of C3 mRNA expression was 8/10 (80%) and 10/13 (77%) in G1 and G2, respectively. The C3 mRNA expression levels were significantly different between the G1 and G2 groups (P = 0.001 and P = 0.01, respectively) (Fig. 3A-B).-

**Fig. 3.**
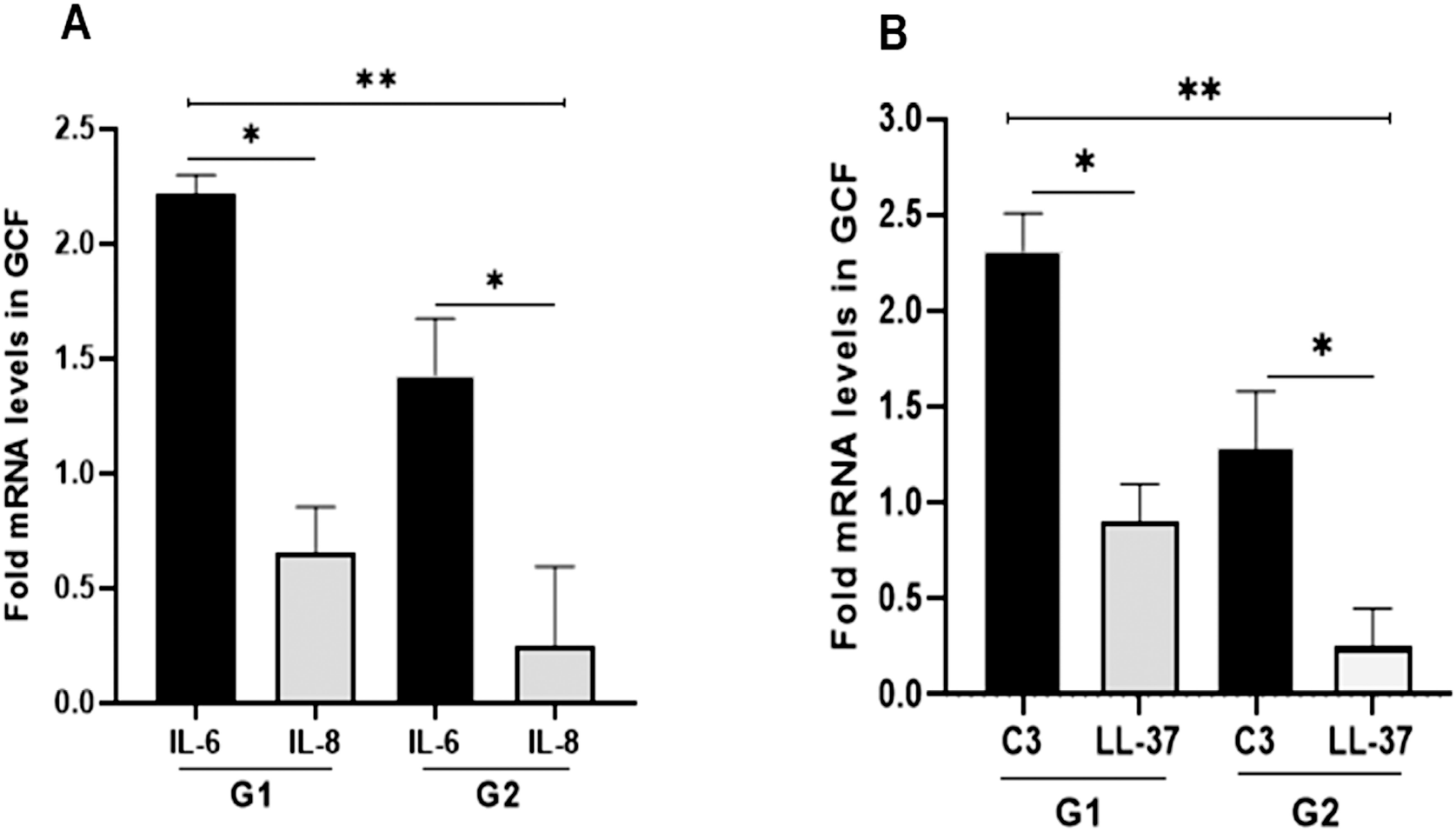
Transcription levels of inflammation determinant genes in gingival crevicular fluids (CGF). To assess inflammation in periodontal tissue, the mRNA expression of IL-6 and IL-8 (A), C3, and LL-37 (B), was measured by qPCR. Data represent mean ± SE of the number of COVID-19 patient separated in two groups: with (G1) and without (G2) diabetes. *P < 0.05 within group, **P< 0.05 between group.

Next, we identified correlations between the transcription levels of IL-6/ IL-8 genes and *ACE2* mRNA expression and the proportion of *P. gingivalis*. In the G1 group, a strong positive and significant correlation was observed between the mRNA levels of the inflammatory cytokine IL-6 and *ACE2* (r = 0.67, P = 0.003) and *P. gingivalis* (r = 0.098, P < 0.0001). In the G2 group, a negative correlation was found between IL-6 and *ACE2* mRNA levels (r = □0.3, P = 0.4), while the correlation between IL-6 mRNA and the relative abundance of *P. gingivalis* was positive (r = 0.92, P < 0.0007) (Fig. 4A-D).

**Fig. 4.**
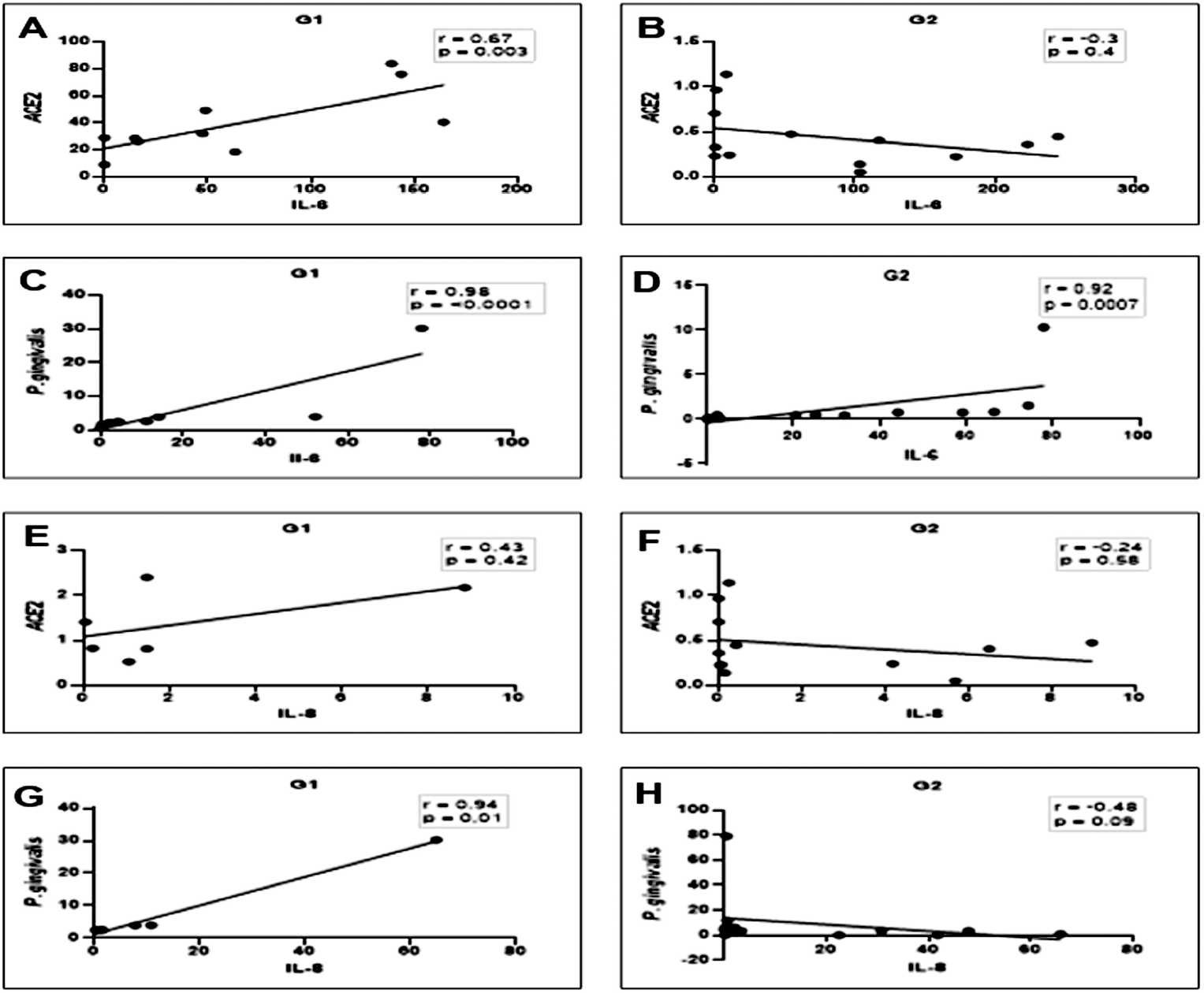
Scatter diagram illustrating the correlation between the mRNA expression of *ACE2* or the proportion of *P. gingivalis* and IL-6/IL8 in periodontitis-COVID-19 patients with (G1) and without (G2) diabetes. These observations indicate that in G1 group, both *ACE2* mRNA transcription and *P. gingivalis* abundance shows strong positive correlation and significant with the transcription levels of IL-6 (A and C). A similar trend was observed in G2 group. The associations between IL-8- and *ACE2* mRNA were moderate and not significant (E) and stronger for *P. gingivalis* (G). In G2 group, the strong significant positive correlation was only observed between *P. gingivalis* and IL-6 (D), while it was negative and for the others (B, F, and H).

For IL-8, a moderate positive correlation was found between *ACE2* mRNA and IL-8 gene expression in the G1 group, however, the association was not significant (r = 0.43 and P = 0.42), In the G1 group, a strong and significant positive correlation was observed between the proportion of *P. gingivalis* and IL-8 mRNA transcription (r = 0.94, P = 0.01). In the G2 group, a negative correlation was found between *ACE2* and IL-8 mRNA levels (r = □0.24, P = 0.58). A similar relationship was found between the IL-8 mRNA levels and *P. gingivalis* infection (r = □048, P = 0.09). Statistically, in the G2 group, neither correlation was significant (Fig. 4A–H).

Correlation analyses were performed to determine any correlations between the transcription level of *ACE2* mRNA and *P. gingivalis* number, and the mRNA transcription of C3 and LL-37, separately for both G1 and G2 groups. The correlation coefficients (r) for all molecules tested (both G1 and G2) are shown in Fig. 5E-H. A strong positive correlation was detected between *ACE2* mRNA and transcription levels of C3 in G1 (r = 0.95, P = 0.01) and G2 (r = 0.95, P = 0.0001). A linear positive correlation was also observed in the G1 group with respect to the correlation between *the number of P. gingivalis* and transcription levels of C3 (r = 0.71, P = 0.008). In G1, the transcription level of C3 mRNA was positively correlated with both *ACE2* mRNA (r = 0.95, P = 0.0001) and the proportion of *P. gingivalis* (r = 0.96, P = 0.006). Lastly, the relative expression of *ACE2* mRNA was strongly and positively correlated with LL-37 transcription in G1 (r = 0.89, P = 0.001) and negatively correlated in G2 (r = - 0.02, P = 0.6); all correlations were not significant (Fig. 5E and F). In contrast, the association between *P. gingivalis* abundance and LL-37 mRNA was strongly positive in both the G1 and G2 groups (r = 0.9, P = 0.001). By referring to the results, we evaluated the accuracy of the combination of IL-6 and C3 mRNAs based on the ROC curve and observed an AUC value of 0.787 for G1 group and 0.55 for G2 group, respectively (Fig. 6A and B).

**Fig. 5.**
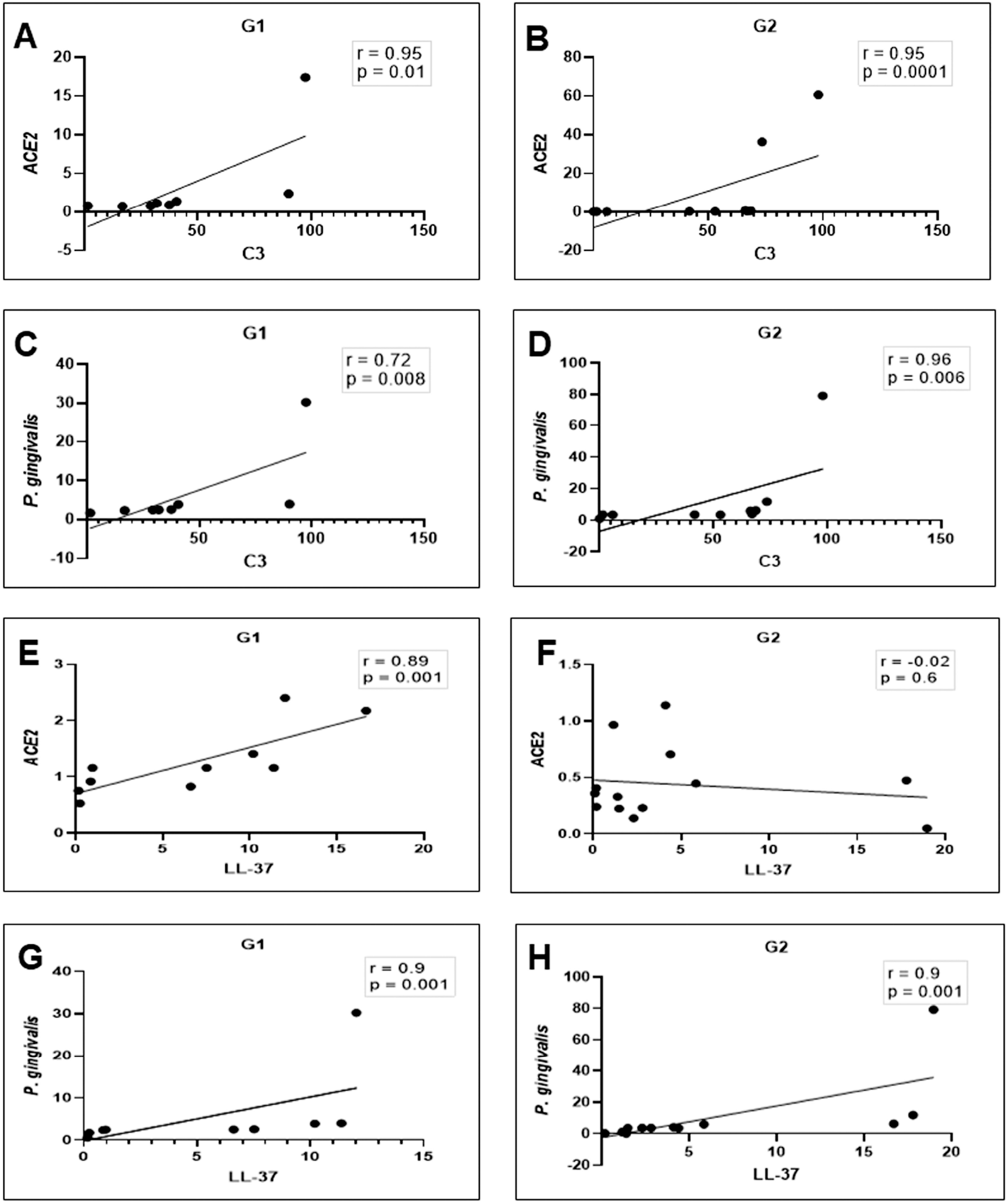
Scatter diagram illustrating the correlation between mRNA expression of *ACE2* or the proportion of *P. gingivalis* and C3/LL-37 in periodontitis-COVID-19 patients with (G1) and without (G2) diabetes. These observations indicate that in G1 group, both the transcription *ACE2* mRNA and *P. gingivalis* abundance show a strong positive correlation and significant with the transcription levels of C3 mRNA and LL-37 (A, C, E, and G). In G2 group, the associations between C3 and *ACE2* mRNA or *P. gingivalis* abundance were strong and significant (A and D), while the correlation with LL-37 was found to be negative not significant between *ACE2*, but positive and significant with LL-37 mRNA expression.

**Fig. 6.**
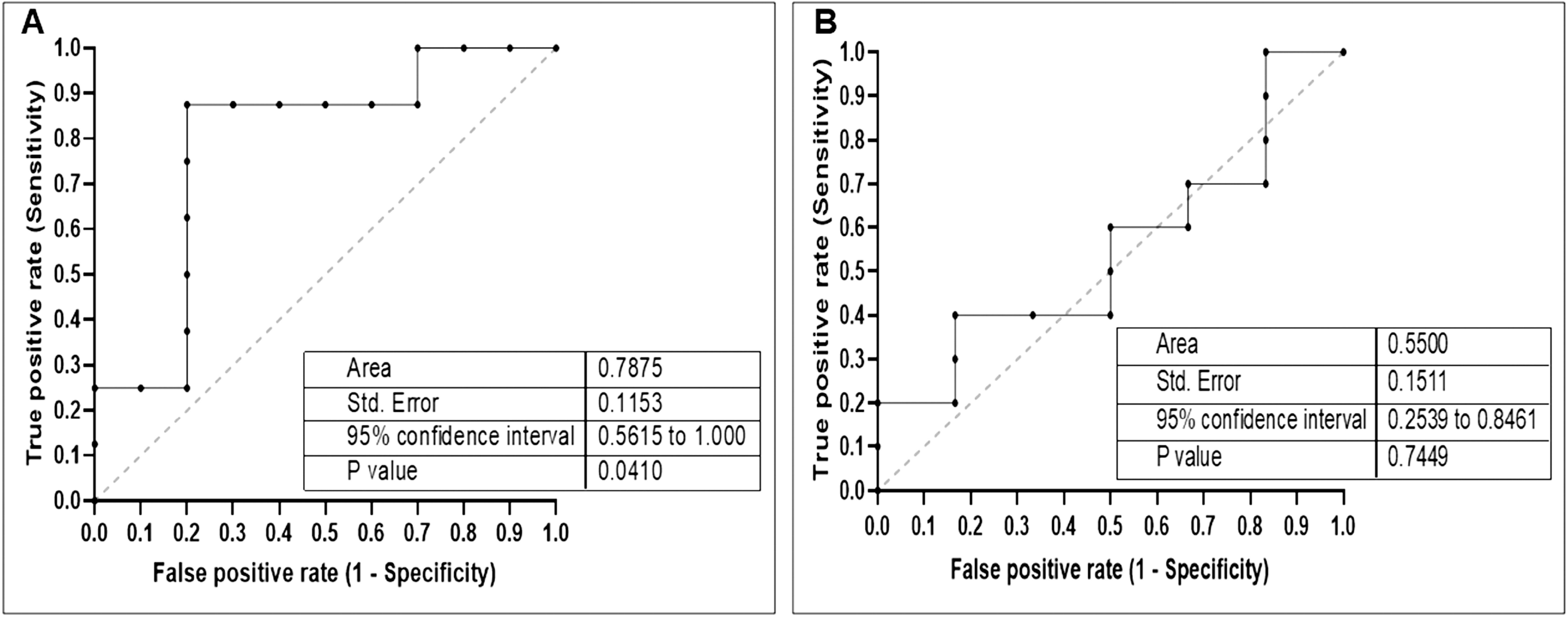
Receiver operating characteristic curve (ROC) showing the plot and the best cut-off point of the relationship between IL-6 and C3. The combination of IL-6 and C3 mRNA expression could discriminate the aggressive of periodontitis in COVID-19 patient with (A) (area under curve/AUC, 0.78; p = 0.04) and without (B) (area under curve/ AUC, 0.5; p = 0.7) diabetes.

## Discussion

In the present study, we used GCF as an oral fluid sample because its components reliably reflect both periodontal and systemic conditions [29, 30]. We assumed that periodontitis and diabetes occurred in patients with COVID-19 prior to COVID-19 infection. Thus, the increased expression level of *ACE2* may indicate that people living with pre-existing chronic conditions of periodontitis with comorbidity (diabetes) have increased vulnerability to SARS-CoV-2 binding by favoring *ACE2* expression in periodontal tissue injury.

Based on this assumption, we first provided evidence that the expression of the functional receptor for SARS-CoV-2-associated molecule (ACE2) can be detected in the subgingival niche of all subjects included in this study. This result is consistent with data showing that the oral mucosa also expresses *ACE2*, especially in tongue epithelial cells [18, 31], which we used to confirm the mRNA expression of *ACE2* in the subgingival habitat. This result may indicate that, although SARS-CoV-2 is a respiratory virus, our study provides evidence that it may invade periodontal tissue, leading to the unique characteristics of the periodontal microenvironment. Additionally, diabetes as a comorbidity condition that accompanies our subjects appears to accelerate the expression of *ACE2* mRNA in the periodontal pocket niche. Since a relationship exists between *ACE2* expression and vulnerability to SARS-CoV-2 infection [32], it is likely that the periodontal tissue of our participants was exposed to SARS-CoV-2 infection. This result could be linked to the clinical status of our COVID-19 patients, which, according to the medical records, showed mild to moderate symptoms (data not shown).

Subsequently, we evaluated the proportion of *P. gingivalis* in COVID-19 patients. Using qPCR, we amplified the 16SrRNA gene region belonging to *P. gingivalis, F. nucleatum*, and *V. parvula*. The three species were detected in the periodontal niche of all subjects included in this study (G1, G2, and control), suggesting their existence as normal oral flora, as well as their association with disease processes in our COVID-19 patients. This result agrees with our previous findings in a subgingival bacterial microbiome diversity study in diabetic patients [26]. In general, the current study found that the most abundant bacteria in both diseased groups (G1 and G2) were *V. parvula*, followed by *F. nucleatum*, while *P. gingivalis* had the lowest abundance. The lowest bacterial abundance indicates its contribution as the key stone species [33] and has an important role in periodontitis and the associated systemic conditions [34-36]. Moreover, as stated by other reports, *P. gingivalis* is a keystone pathogen capable of subverting inflammatory and innate responses [37], while *F. nucleatum* and *V. parvula* are two crucial species, as they play an important role in shaping oral biofilm ecology [38-40]. These two bacteria are positively related to COVID-19-associated events [41, 42]. Our observations suggest that the relationships between the three bacteria may reflect the characteristics of the subgingival microbiome in periodontitis patients with diabetes [26], and except for *V. parvula* (as shown in Fig. 1B), the diabetes condition did not modulate the proportion pattern shown by *P. gingivalis* and *F. nucleatum* in the periodontal niche of COVID-19 patients. Additionally, we found that in both investigated groups, the lowest abundance of *P. gingivalis* was positively correlated with relatively higher propositions of *F. nucleatum* and *V. parvula*.

This association indicates that, as a keystone pathogen, *P. gingivalis* requires an additional oral microbiota to induce periodontitis [43]. Thus, by referring to the positive relationships between the *P. gingivalis* proportion and the expression of *ACE2* mRNA, it is likely that the aggressive pathogenic subgingival flora in our COVID-19 patients may also include SARS-CoV-2, since the virus can be detected in GCF [11], and the role of *P. gingivalis* in accelerating the expression level of *ACE2* has been reported in in vitro studies [44]. In addition, our results confirmed the results of an *in vitro* study, which reported on the upregulation of *ACE2* in alveolar epithelial cells after exposure to *F. nucleatum* [41]. In the present study, it was not possible to determine whether the proportion of the species observed in the subgingival microbial community was the cause or effect of SARS-CoV-2 replication or pathogenesis. However, the low expression of *ACE2* in GCF (compared to TS) and the proportion of *P. gingivalis* may explain the presence of SARS-CoV-2 in the periodontal pocket microenvironment, favoring the emergence and persistence of dysbiosis in periodontitis. Further studies will be necessary to address this hypothesis.

The key stone pathogen (*P. gingivalis*) and other dysbiotic-related bacteria are the initiators and progression of periodontitis. Its low proportion would more likely capture a state of periodontitis activity in our subjects, which is compatible with the existence of infection with associated inflammatory responses. In this study, we detected the levels of mRNA expression of four selected innate immunity markers (IL-6, IL-8, C3, and LL-37) using qPCR in patient groups that were separated based on diabetes status. We did not include other comorbidities; therefore, we were not able to correlate and possibly distinguish the levels of the tested biomarker for another concomitant medical comorbidity. Remarkably, we found that IL-6 and C3 were detected at high (>2-fold) and intermediate (>1-fold) levels in the GCF of COVID-19 patients with and without diabetes, respectively. Additionally, we observed the same trend in the transcription levels of IL-6 vs. IL-8 and C3 vs. LL-37 in both patient groups, where the mRNA expression of IL-6 and C3 was generally higher in the G1 group (COVID-19 patients with diabetes). The upregulation of IL-6 is in line with the higher proportion of *V. parvula*, as this species has been shown to induce the production of the pro-inflammatory cytokine (IL-6) [45]. Furthermore, the transcription levels of *ACE2* and the abundance of *P. gingivalis* were positively correlated with low levels of IL-8 or LL-37 mRNA expression. This association may indicate the severity of bacterial infection in periodontal habitats [46-48], particularly in COVID-19 patients with diabetes, where SARS-CoV-2 and *P. gingivalis* may paralyze the chemokine (IL-8) locally induced by the presence of mixed bacteria [49], represented by *F. nucleatus* and *V. parvula*. Indeed, our data further indicated a more severe bacterial infection of periodontitis, as indicated by the lower expression of antimicrobial peptide (LL-37 mRNA).

During COVID-19 infection, it is difficult to assess the periodontal status that is normally supported by radiographic evidence to clarify the bone level, and pathological features are key parameters in such situations. In this sense, the cytokine IL-6 is a biomarker in people with periodontal disease [50, 51], whereas complement C3 is a crucial inflammatory element that contributes to the persistence of infection in the microbial community, as its involvement leads to a destructive inflammatory response in periodontal tissue [52, 53]. Our observations provide additional information, in which there is a strong correlation between dysbiotic-related key stone bacterium (*P. gingivalis*) and the amplification of inflammation in the periodontal niche, as indicated by the upregulation of IL-6 and C3 mRNA expression.

We noted that the transcription levels of both IL-6 and C3 in the periodontal niche were positively associated with the upregulation of *ACE2* mRNA and relatively lower abundance of *P. gingivalis* in the periodontal microenvironment of the G1 group. If the transcription levels of *ACE2* were positively associated with the expression of the related proteins, our data suggest that the upregulation of *ACE2* in a dysbiosis microenvironment (periodontal niches) functions in response to the presence of SARS-CoV-2, leading to cellular entry of the virus and acceleration of the secretion of inflammatory mediators, including IL-6 and C3, as we observed in this study. This may affect the existence of periodontal inflammation and the survival of the bacterial community in the subgingival area (represented by the three periodontal pathogens), which initiates inflammation. A similar biological phenomenon has been reported in gut microbiome dysbiosis [54]. Taken together, these findings suggest that as the transcription levels of both cytokines in the periodontal microenvironment of the G1 group (COVID-19 patient with diabetes) had a positive association with the upregulation of *ACE2*. During the hospital stay, the systemic condition of our patients may be involved in the regulation of the proinflammatory marker. Hence, higher transcription levels of inflammatory markers (IL-6 and C3) may play an important role in stabilizing the inflammatory process, both locally and systemically [29, 55]. However, the presence of SARS-CoV-2 in periodontal compartments, which contain pathogenic bacteria, is assumed to be indirectly involved in the destruction of periodontal tissue. Although in the current study we could not evaluate the effect of these biological markers as pathogenic or protective-related mediators in our COVID-19 subjects, it is likely that IL-6 and C3 could be comparable to predict the activity of periodontal disease in patients who have been infected with SARS-CoV-2. Therefore, to improve the prediction accuracy of the combination of IL-6 and C3 markers in G1 and G2 groups, it is necessary to identify the sensitivity and specificity to obtain the most informative features. Hence, based on ROC curve analyses, we found that both IL-6 and C3 mRNA expression had a higher predictive value for periodontitis activity in COVID-19 patients with diabetes. Since IL-6 levels have been reported to be a very important predictor of severe COVID-19 infection [56-58], we suggest that there is synergistic activity between IL-6 and complement C3 as the underlying mechanism of periodontal disease activity in COVID-19 patients with diabetes. Thus, complement C3 may serve as a potential biomarker of periodontal inflammation and disease severity [59] in COVID-19 patients with diabetes. These results indicated that the complement C3 can serve as a potential biomarker of periodontal inflammation and disease severity in COVId-19 patient with diabetes with certain degree of accuracy. More studies are necessary to address a such assumption.

Our study has intrinsic limitations. First, as an observational study, it was difficult to exclude all potential confounders. Therefore, the involvement of some variables may be underestimated in predicting the result. Second, the number of subjects involved was small, and we could not be sure if the altered microbial composition in the periodontal niche had already occurred in our COVID-19 patients when GCF samples were collected. Periodontitis is a common infectious disease that is triggered by oral bacteria. It is important to analyze certain quantities of specific pathogens in the oral microbiota of patients to initiate inflammation. In this study, we used the relative abundance of each targeted species as a proportion rather than the actual levels. Finally, although GCF reflects the serum immune response [11], the results of this study cannot be linked directly to suggest that the level of inflammation determinants in GCF may correspond to the systemic inflammation mechanism of COVID-19. Based on the results of this preliminary study, we are planning a forthcoming study in searching the oral microbiome and trying to correlate the viral load and the functional inference of oral bacterial composition, and how it relates to the host response. However, with these limitations, the current approach hints at the importance of the oral ecosystem in modulating clinical symptoms related to COVID-19 cases [60-63].

## Conclusion

Within the limitations of the present study, our data revealed that the periodontitis microenvironment in COVID-19 patients with diabetes maintains a distinctive relationship between SARS-CoV-2 entry molecules and dysbiotic conditions in the periodontal microenvironment. This relationship could be associated with the bacterial composition due to the presence of the virus, but the involvement of comorbidity (diabetes) needs to be considered. The results of this preliminary study could help to understand the underlying association between COVID-19 and periodontal disease, and its relationship with periodontopathogens represented by *P. gingivalis, F. nucleatum*, and *V. parvula* in the inflammation process, which could be used to distinguish between active and non-active periodontitis conditions during the monitoring of the medical condition of COVID-19 patients.

## Supporting information

https://drive.google.com/drive/folders/17GyE-3Ew0pBvsMqY4UusQB2SCdsVdcVL?usp=sharing

## Data Availability

The data underlying the results presented in the study are available from the corresponding author. Please send email to

## Acknowledgements

All the authors would like to acknowledge all the study participants, Director of RSUI and the Ethical clearance committee for providing permission and ethical clearance to conduct this research project. This study was supported by The Grant from Ministry of Education and Culture of the Republic of Indonesia. (No.NKB-143/UN2.RST/HKP.05.00/2021).

## Author Contribution

**Conceptualization**: BM. Bachtiar, EW. Bachtiar, Y. Soeroso, H. Sunarto

**Data Curation**: Y. Soeroso, H. Sunarto, B. Sulijaya, IP. Pratomo, A. Kusumaningrum, Yudhistira, CF.Theodorea, E. Apriyanti, D. Efendi.

**Formal analysis**: BM.Bachtiar, EW.Bachtiar

**Clinical Investigation**: IP. Pratomo, A. Kusumaningrum, Yudhistira, CF. Theodorea, B. Sulijaya, E. Apriyanti, D. Efendi.

**Methodology**: BM.Bachtiar, Y. Soeroso, H. Sunarto, B. Sulijaya

**Supervision-Clinical-Lab**: IP. Pratomo, A. Kusumaningrum, Yudhistira.

**Technical Lab**: NR. Utami, A. Deviana, AD. Andiryani

**Writing-original draft**: BM. Bachtiar

**Writing-review &editing**: EW. Bachtiar

## Data Availability

The data that support the findings of this study are available from Oral Science Research Center, Faculty of Dentistry Universitas Indonesia, Indonesia. Data are available from the corresponding authors upon reasonable request and permission of Dental Center Universitas Indonesia Hospital.

